# A systematic review and meta-analysis of the diagnostic accuracy of endometrial sampling tests for detecting endometrial cancer

**DOI:** 10.1101/2023.01.18.23284733

**Authors:** Noha Sakna, Marwa Elgendi, Mohamed Salama, Ahmed Zeinhom, Somia Labib, Ashraf Nabhan

## Abstract

**Objectives:** to determine the diagnostic accuracy of different endometrial sampling tests for detecting endometrial carcinoma.

**Design:** a systematic review and meta-analysis of studies of diagnostic accuracy.

**Eligibility criteria:** We included published diagnostic test accuracy studies of women, of all ages, who had an endometrial sampling for preoperatively detecting endometrial cancer with verification using histopathology of hysterectomy specimens as the reference standard. We excluded case control and case series studies.

**Information sources:** We searched the Cochrane library, MEDLINE/PubMed, CINAHL, Web of Science, and Scopus from the date of inception of the databases to January 18, 2023. We did not apply any restrictions on language or date of publication. We searched the references of included studies and other systematic reviews.

**Risk of bias:** We extracted study data and assessed study quality using the revised quality assessment tool for diagnostic accuracy studies (QUADAS-2).

**Synthesis of results:** We used bivariate diagnostic random-effects meta-analysis and presented the results in a summary receiver operating characteristic curve. We assessed the certainty of evidence as recommended by the GRADE approach.

**Results:** Twelve included studies, published between 1986 and 2022, recruited 1607 participants. Seven studies were low risk of bias in all domains and all studies had low applicability concerns. The most examined index tests were Pipelle and conventional dilation and curettage. For diagnosing endometrial carcinoma, the sensitivity, specificity, positive likelihood ratio, and negative likelihood ratio (95% confidence intervals), for Pipelle were 0.774 (0.565, 0.900), 0.985 (0.927, 0.997), 97.000 (14.000, 349.000), and 0.241 (0.101, 0.442)and for conventional dilation and curettage were 0.773 (0.333, 0.959), 0.987 (0.967, 0.995), 62.300 (18.600, 148.000), and 0.268 (0.042, 0.676); respectively.

**Conclusion:** High certainty evidence indicates that pre-operative endometrial sampling particularly using Pipelle or conventional curettage is accurate in diagnosing endometrial cancer. Studies assessing other endometrial sampling tests were sparse.

**Systematic review registration:** Center for Open Science, osf.io/h8e9z

## Introduction

Endometrial cancer (EC) is the fifth most common malignant condition among women worldwide. Its incidence shows an increase in line with an increase in life expectancy. It is a major health concern being the 14th leading cause of death from cancer in women.^1–5^

Most cases affect postmenopausal women^6^. Most women present with abnormal uterine bleeding.^7^ While 90 percent of women with endometrial cancer initially present with PMB, only 5 to 10 percent of women with PMB are diagnosed with endometrial cancer. Irregular pre- or peri-menopausal bleeding is yet another important presenting symptom.^7^ Endometrial cancer is sometimes diagnosed in women without an abnormal uterine bleeding. This occurs during the investigation of a thick endometrial line found on an imaging technique performed for other reasons or incidentally in hysterectomy specimens performed for benign conditions.^4,8^

Survival from endometrial cancer depends on the stage at diagnosis. By enabling an early detection of atypical hyperplasia and early-stage endometrial cancer, an accurate early detection remains the cornerstone for improving outcomes.^4,9,10^

### Objectives

The aim of this systematic review and meta-analysis is to determine the diagnostic accuracy of different endometrial sampling tests for detecting endometrial carcinoma.

## Methods

We conducted this systematic review using methodological approaches prespecified in a review protocol ^11^ which was prospectively registered in the Center for Open Science. ^12^ We reported this review according to the Preferred Reporting Items for a Systematic Review and Meta-analysis of Diagnostic Test Accuracy Studies (PRISMA-DTA) guidelines. ^13^

### Eligibility criteria

We included published a one-gate design studies that evaluated the diagnostic accuracy of different endometrial sampling methods, followed by verification of study participants with a reference test. Participants are women, of all ages, who had an endometrial sampling for a suspected endometrial hyperplasia or cancer. Index tests included different endometrial sampling techniques. Target conditions included endometrial hyperplasia or carcinoma. We accepted histopathology of hysterectomy specimens as the reference standard for the diagnosis of the target conditions. We excluded studies of a case-control design that included women with known endometrial carcinoma to matched controls.^14^

### Information sources

We developed a comprehensive search strategy to find published articles.^15^ We searched the Cochrane library, MEDLINE/PubMed, CINAHL, Web of Science, and Scopus from the date of inception of the databases to January 18, 2023. We did not apply any restrictions on language or date of publication.

#### Searching other resources

We manually searched the reference lists in articles retrieved from electronic databases and relevant review articles.

### Study selection

Two review authors (SL and NS) used a reference manager, to import and de-duplicate search results. They independently screened titles and abstracts to identify potentially eligible studies for full-text retrieval. Two review authors (SL and MA) double checked the list of potentially eligible studies. Review authors working in pairs, independently assessed the full text using the predefined eligibility criteria. They resolved discrepancies by discussion and by consulting with the lead author (AN). We illustrated the study selection process using a PRISMA flowchart.

### Data collection process

For each included study, two review authors (SL, MA), independently extracted the following data: General information: title, journal, year, and study design; Sample size: total number of participants included and tested; Baseline characteristics: age, index test, reference test; numbers of true positive, true negative, false positive, and false negative findings. We extracted these data for both target conditions (AEH and EC). All numbers were double checked by two authors (NS, AZ). We summarized the data from each study in 2 × 2 tables (true positive, false positive, false negative, true negative), and we stored the data into a spreadsheet.

### Risk of bias and applicability

Review authors, working in pairs, independently assessed the risk of bias of included studies and applicability of their results using QUADAS-2. ^16^ They resolved discrepancies by discussion and by consulting with the lead author. We addressed aspects of study quality involving the participant spectrum, index tests, target conditions, reference standards, and flow and timing. ^16^ We classified a study as having a high risk of bias if at least one of the domains of QUADAS-2 was judged as being high risk.

### Synthesis of results

Each method of endometrial sampling was compared against the reference test. For each method of endometrial sampling, TP, TN, FP, and FN were computed. We used bivariate diagnostic random-effects meta-analysis. ^17^ We presented the results in a summary receiver operating characteristic (ROC) curve. We used the Markov chain Monte Carlo procedure to generate summary positive and negative likelihood ratio and diagnostic odds ratio for the bivariate model. ^18^ When there are three or less studies, we used a univariate random-effects model. For each of the target condition, we presented the synthesis by the method of sample collection. We assessed the effect of risk of bias of included studies on diagnostic accuracy by performing a sensitivity analysis in which we excluded studies classified as having high risk of bias. We used R software version 4.2.2 ^19^ and package ‘mada’. ^20^

### Summary of findings table and assessment of the certainty of evidence

We prepared summary of findings tables, using GRADEpro GDT^21^, to present the main results and key information regarding the certainty of evidence. We assessed the certainty of evidence as recommended by the GRADE approach.^22^

We rated the certainty of evidence as either high (when not downgraded), moderate (when downgraded by one level), low (when downgraded by two levels), or very low (when downgraded by more than two levels) based on five domains: risk of bias, indirectness, inconsistency, imprecision, and publication bias. For each outcome, the certainty of evidence started as high when there were high-quality observational studies (cross-sectional or cohort studies) that enrolled participants with diagnostic uncertainty. If we found a reason for downgrading, we used our judgement to classify the reason as either serious (downgraded by one level) or very serious (downgraded by two levels).^23,24^

### Patient and public involvement

Patients and/or the public were not involved in the design, or conduct, or reporting, or dissemination plans of this research.

## Results

### Study selection

Bibliographic database search identified 3,386 records and 23 additional records from searching the reference lists of relevant records. After removing duplicates, 2679 titles and abstracts were screened. Full text reports of 38 potentially eligible studies were assessed using the predefined eligibility criteria and twelve studies were included, Figure 1.

**Figure 1.**
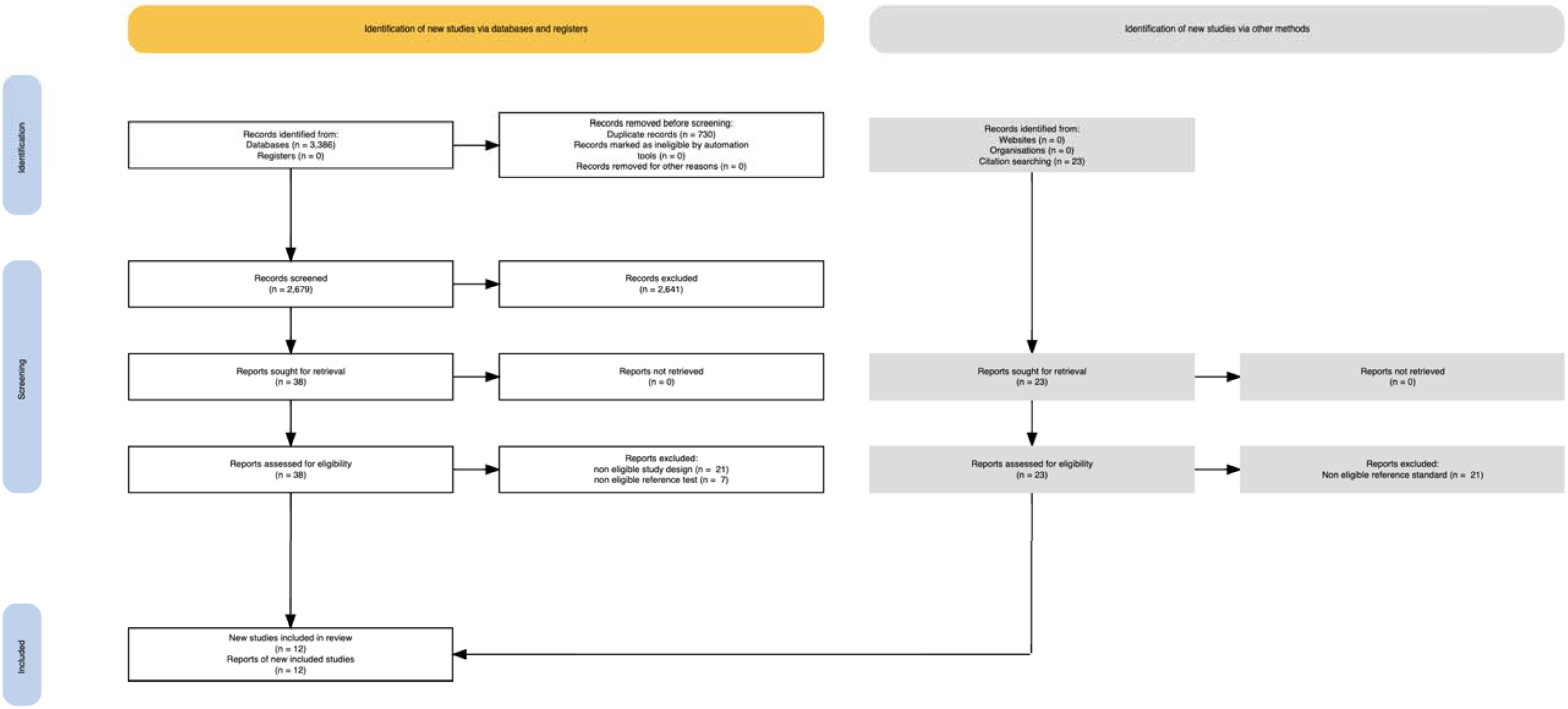
PRISMA Flow Diagram.

### Study characteristics

The twelve included studies, published between 1986 and 2022, had a one-gate design and recruited 1607 participants, Table 1.

**Table 1:** Characteristics of included studies

| Author | Year | Design | Outcomes | Sample size | Index test | Reference test | Participants |
| --- | --- | --- | --- | --- | --- | --- | --- |
| Behnamfar<sup>25</sup> | 2020 | A one-gate diagnostic test accuracy | AEH, EC | 25 | Pipelle, D&C | Histopathology examination of hysterectomy | All women were menopausal, with post-menopausal bleeding (mean age 60) |
| de Leon<sup>28</sup> | 2016 | A one-gate diagnostic test accuracy | AEH, EC | 42 | GDP Tao device | Histopathology examination of hysterectomy | Pre- and post-menopausal women, mostly with abnormal uterine bleeding (median age 57) |
| Gerretsen<sup>29</sup> | 1987 | A one-gate diagnostic test accuracy | AEH, EC | 61 | Abradul cell sampler | Histopathology examination of hysterectomy | Pre- and post-menopausal women who had a total hysterectomy |
| Gungorduk<sup>26</sup> | 2014 | A one-gate diagnostic test accuracy | AEH, EC | 189 | Pipelle, D&C | Histopathology examination of hysterectomy | Pre- and post-menopausal women who had a total hysterectomy (mean age 49) |
| Han<sup>30</sup> | 2019 | A one-gate diagnostic test accuracy | AEH, EC | 271 | Li brush | Histopathology examination of hysterectomy | Pre- and post-menopausal women who had a total hysterectomy |
| Ilavarasi<sup>27</sup> | 2019 | A one-gate diagnostic test accuracy | AEH, EC | 106 | Pipelle | Histopathology examination of hysterectomy | Pre- and post-menopausal women with AUB |
| Kufahl<sup>31</sup> | 1997 | A one-gate diagnostic test accuracy | AEH, EC | 181 | Gynoscann, UEC | Histopathology examination of hysterectomy | Pre- and post-menopausal women who had a total hysterectomy (median age 48) |
| Moradan<sup>35</sup> | 2017 | A one-gate diagnostic test accuracy | EC | 163 | D&C | Histopathology examination of hysterectomy | Pre- and post-menopausal women with AUB resistant to medical therapy, or AUB in combination with an endometrial thickness of more than 12 mm on performing transvaginal ultrasound |
| Reddington<sup>33</sup> | 1995 | A one-gate diagnostic test accuracy | AEH, EC | 25 | Masterson curette | Histopathology examination of hysterectomy | Pre- and post-menopausal women who had a total hysterectomy |
| Saadia<sup>34</sup> | 2011 | A one-gate diagnostic test accuracy | AEH, EC | 50 | D&C | Histopathology examination of hysterectomy | Pre- and post-menopausal women who had a total hysterectomy (median age 43) |
| Somasundar<sup>36</sup> | 2022 | A one-gate diagnostic test accuracy | EC | 54 | D&C, Hysteroscopy | Histopathology examination of hysterectomy | Pre- and post-menopausal women with AUB |
| Stever<sup>32</sup> | 1986 | A one-gate diagnostic test accuracy | AEH, EC | 440 | Novak | Histopathology examination of hysterectomy | Pre- and post-menopausal women who had a total hysterectomy (mean age 40) |

The majority of recruited women were postmenopausal. Participants had different presentations including post menopausal bleeding, pre-menopausal abnormal uterine bleeding, or women scheduled for hysterectomy for different indications.

Studies used different methods of endometrial sampling. This included Pipelle^25–27^, GDP Tao^28^, Abradul^29^, Li brush^30^, Uterine Explora Curette (UEC)^31^, Gynoscann^31^, Novak^32^, M curette^33^, or D&C.^25,26,34^ The search did not identify an eligible study, as per the presepcified criteria for this review, that used an outpatient hysteroscopy-directed endometrial sampling. All studies used histopathology examination of hysterectomy as the reference standard. All studies reported EC and AEH.

### Risk of bias and concerns regarding applicability

Regarding bias, seven studies were classified as low risk in all domains. Two studies had a high risk of bias, one in the domain of flow and timing in one study and one in the reference standard due to partial verification. Four studies had an unclear risk of bias in the domains of patient selection, reference standard, and flow and timing. All studies had low concerns regarding applicability, Table 2.

**Table 2:** Quality assessment of diagnostic accuracy studies (QUADAS-2)

| Study ID | Risk of bias |  |  |  | Concerns regarding applicability |  |  |
| --- | --- | --- | --- | --- | --- | --- | --- |
|  | Patient selection | Index test | Reference standard | Flow and timing | Patient selection | Index test | Reference standard |
| Behnamfar, 2020 | Unclear | Low | Unclear | High | Low | Low | Low |
| de Leon, 2016 | Low | Low | Low | Low | Low | Low | Low |
| Gerretsen, 1987 | Low | Low | Low | Low | Low | Low | Low |
| Gungorduk, 2014 | Low | Low | Low | Low | Low | Low | Low |
| Han, 2019 | Low | Low | Low | Low | Low | Low | Low |
| Ilavarasi, 2019 | Low | Low | Low | Low | Low | Low | Low |
| Kufahl, 1997 | Low | Low | Low | Low | Low | Low | Low |
| Moradan, 2017 | Low | Low | Low | Unclear | Low | Low | Low |
| Reddington, 1995 | Low | Low | Low | Low | Low | Low | Low |
| Saadia, 2011 | Low | Low | Unclear | Low | Low | Low | Low |
| Stever, 1986 | Low | Low | Low | Unclear | Low | Low | Low |
| Somasundar, 2022 | Low | Low | High | Low | Low | Low | Low |

### Data synthesis

#### Diagnostic accuracy of endometrial sampling tests compared to histopathology of hysterectomy

Twelve studies reported data on endometrial cancer^25–36^ and ten reported data on atypical endometrial hyperplasia using different methods of endometrial sampling, Table 3, Figure 2, and Figure 3.^25–34^

**Table 3:** GRADE Summary of findings table: Diagnostic accuracy of all endometrial sampling tests compared to histopathology of hysterectomy

| Index test | Target condition | Number of studies (participants) | Sensitivity (95% CI) | Specificity (95% CI) | Positive LR (95% CI) | Negative LR (95% CI) | Number of results per 1,000 patients tested (95% CI) Assumed Prevalence 10% for endometrial cancer or atypical hyperplasia |  |  |  | Certainty of the evidence |
| --- | --- | --- | --- | --- | --- | --- | --- | --- | --- | --- | --- |
|  |  |  |  |  |  |  | True positives will receive appropriate treatment | False negatives will be misdiagnosed and not receive appropriate treatment | True negatives will not undergo inappropriate treatment or unnecessary further testing | False positives will undergo inappropriate treatment |  |
| All tests | Endometrial cancer | 12 (1607) | 0.781<br>(0.669, 0.863) | 0.989<br>(0.982, 0.994) | 74.800<br>(42.100, 123.000) | 0.226<br>(0.139, 0.335) | 78 (67 to 86) | 22 (14 to 33) | 890 (884 to 895) | 10 (5 to 16) | ⊕⊕⊕⊕<br>High |
|  | Atypical hyperplasia | 10 (1390) | 0.760<br>(0.639, 0.849) | 0.978<br>(0.953, 0.989) | 36.100<br>(16.400, 69.700) | 0.250<br>(0.155, 0.368) | 76 (64 to 85) | 24 (15 to 36) | 880 (858 to 890) | 20 (10 to 42) | ⊕⊕⊕⊕<br>High |

**Figure 2.**
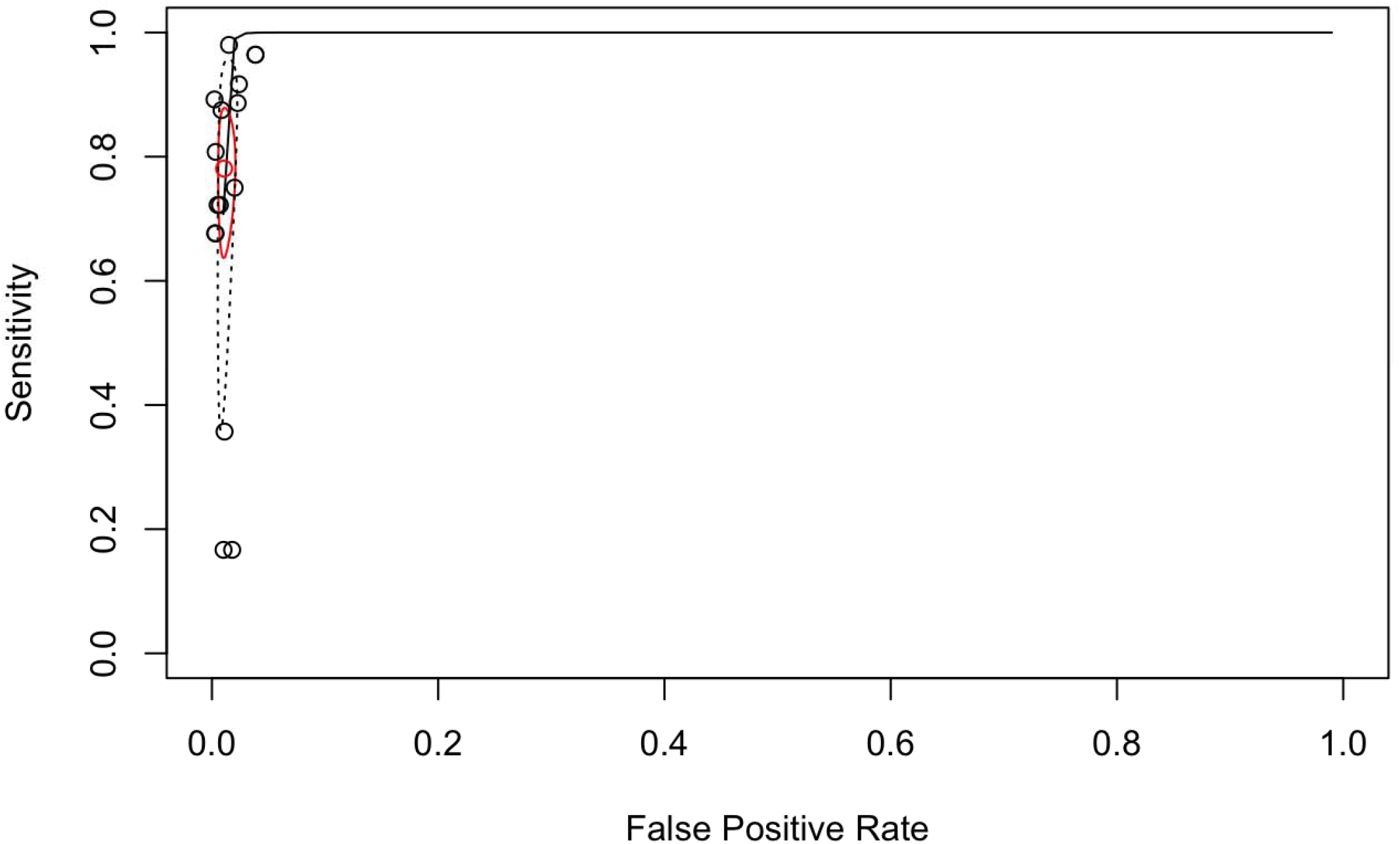
Summary receiver operating characteristic curve for all endometrial sampling tests for detecting endometrial carcinoma.

**Figure 3.**
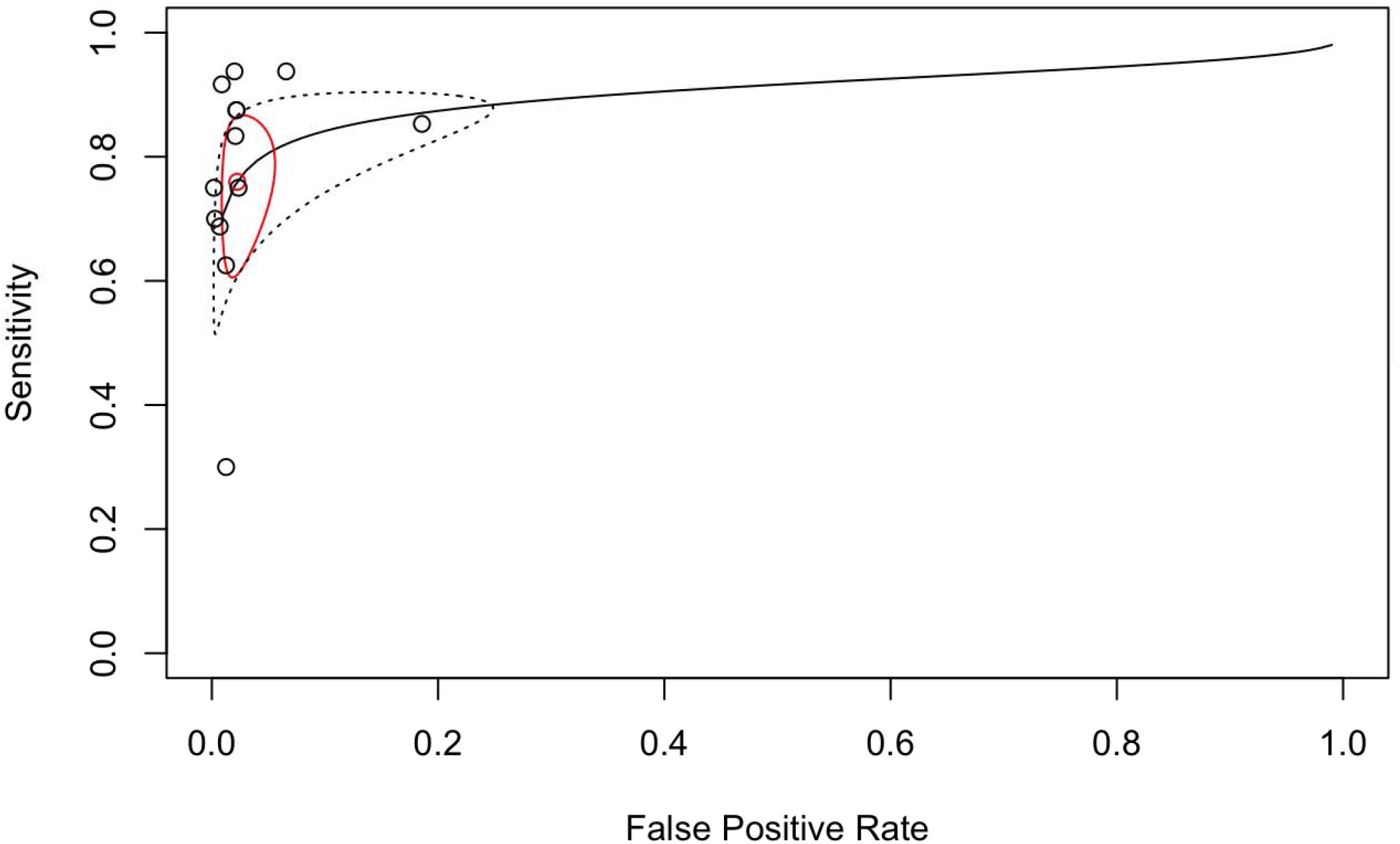
Summary receiver operating characteristic curve for all endometrial sampling tests for detecting atypical endometrial hyperplasia.

For endometrial cancer, out of 100 people with endometrial cancer, 78 would be correctly diagnosed and 22 would be incorrectly diagnosed. Out of 100 people without endometrial cancer, 99 would be correctly diagnosed and 1 would be incorrectly diagnosed. For atypical hyperplasia, out of 100 people with atypical hyperplasia, 76 would be correctly diagnosed and 24 would be incorrectly diagnosed. Out of 100 people without atypical hyperplasia, 98 would be correctly diagnosed and 2 would be incorrectly diagnosed.

#### Subgroup analysis by the specific test

The measures of diagnostic accuracy, sensitivity, specificity, positive LR, and negative LR, for each test are shown in Table 4

**Table 4:**
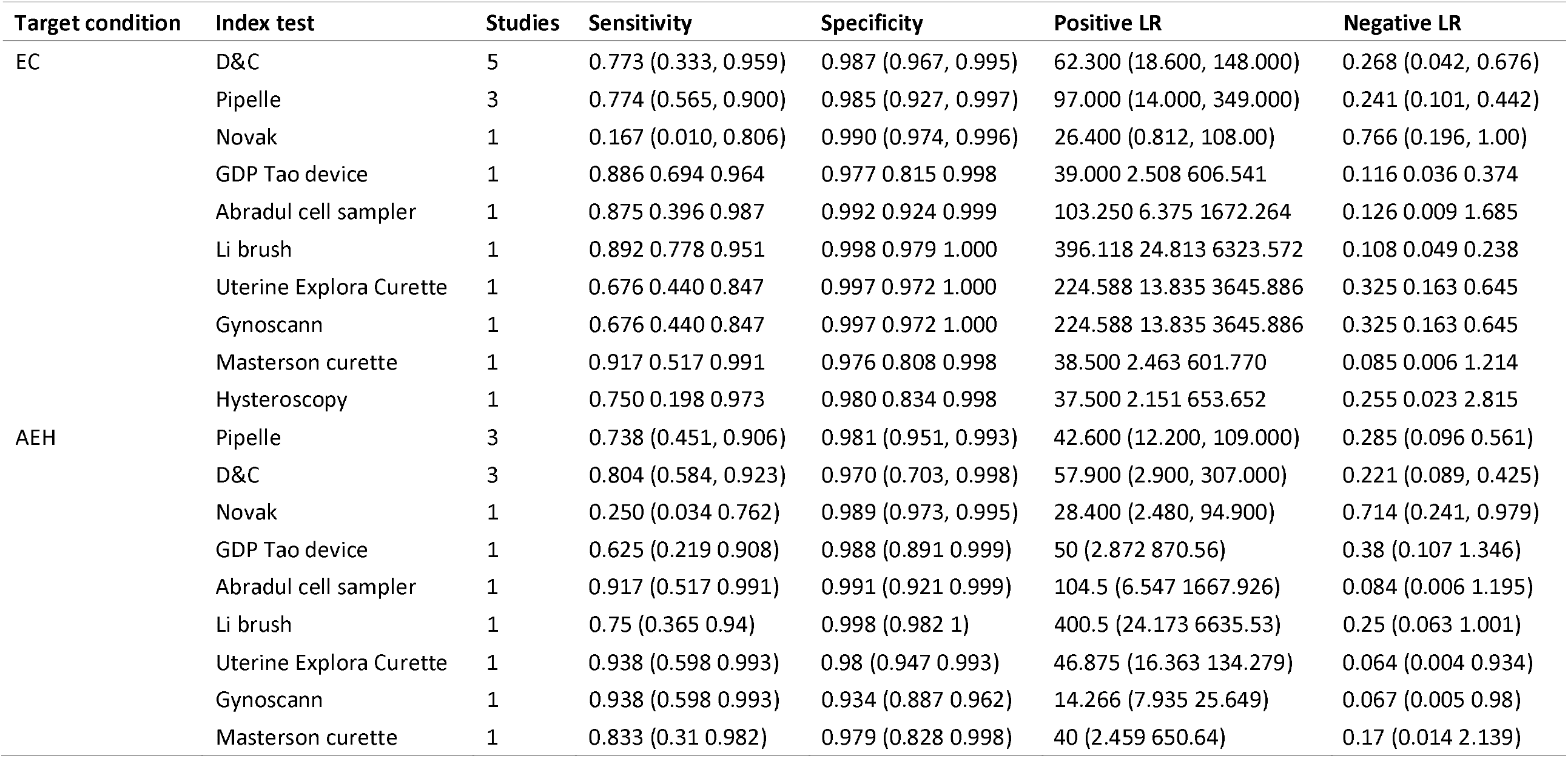
Test accuracy for individual endometrial sampling tests for diagnosing endometrial cancer and atypical endometrial hyperplasia

Five studies examined the accuracy of D&C for diagnosing EC.^25,26,34–36^. Using D&C, compared to histopathology examination of hysterectomy, out of 100 people with endometrial cancer, 77 would be correctly and out of 100 women without endometrial cancer 99 would be correctly diagnosed, Table 4. Three studies reported the accuracy of D&C for diagnosing AEH.^25,26,34^ Using D&C, compared to histopathology examination of hysterectomy, out of 100 people with AEH, 80 would be correctly and out of 100 women without AEH 97 would be correctly diagnosed.

Three studies examined the accuracy of Pipelle for diagnosing AEH or EC.^25–27^ Using Pipelle endometrial sampling, compared to histopathology examination of hysterectomy, out of 100 people with endometrial cancer, 77 would be correctly diagnosed and out of 100 women without endometrial cancer 99 would be correctly diagnosed. Using Pipelle, compared to histopathology examination of hysterectomy, out of 100 people with AEH, 74 would be correctly and out of 100 women without AEH 98 would be correctly diagnosed.

Diagnostic accuracy was examined in a single study for each of the following tests: Novak^32^, GDP Tao device^28^, Abradul cell sampler^29^, Li brush^30^, Uterine Explora Curette^31^, Gynoscann^31^, Masterson Curette^33^, and Hysteroscopy^36^, Table 4.

## Discussion

### Summary of the evidence

This systematic review assessed the diagnostic accuracy of various endometrial sampling tests for detecting endometrial cancer or atypical endometrial hyperplasia. Only studies with a one gate design were considered for inclusion in the review. Twelve DTA studies were identified, including 1607 women. All studies were published in peer-reviewed journals and most of these studies were of high methodological quality according to the QUADAS-2 criteria. Pooled estimates of diagnostic test accuracy were computed for various endometrial sampling tests. Robust evidence based on the results of the present review indicate that using Pipelle or conventional curettage can correctly confirm or exclude endometrial cancer or atypical endometrial hyperplasia. High quality evidence is insufficient for hysteroscopy, GDP Tao device, Abradul cell sampler, Li brush, Uterine Explora Curette, Gynoscann, Z-sampler, Novak, and Masterson curette.

This systematic review represents the synthesis of exclusively high quality diagnostic accuracy studies. A one-gate design studies in patients with diagnostic uncertainty and direct comparison of endometrial sampling test results with the appropriate reference standard (histopathology of hysterectomy) are considered high quality.

Every effort was made to minimize the risk of bias. First, we did not include case control studies or studies that include only women with endometrial carcinoma. Previously published reviews have included studies with case control design (two-gate design) which is generally not representative of a test’s accuracy in clinical practice.^9,37^ Spectrum bias through case-control design would lead to sub-optimal patient selection. This falsely inflates sensitivity and specificity. Further, both prevalence and predictive values depend on the ratio of people with and without the disease. In case–control studies, this ratio is constructed artificially, and thus prevalence and predictive values calculated from such a study are artefacts.^38–40,41,42^ Other reviews have primary included studies of index tests in women with known endometrial carcinoma. The ability of the test to rule out the disease can never be calculated if the study recruited only participants known to have the condition.^42,43^

Second, we only included studies that used histopathology examination of hysterectomy as a reference test. Previous reviews has used multiple verification tests and tests that might not correctly classify the disease.^9,44,45^ Errors in the reference test cause misclassification bias. Misclassification can significantly underestimate sensitivity and specificity. The magnitude of the bias depends on the disease prevalence, the accuracy of the index test, and the degree of misclassification. The misclassification rate can vary from study to study depending on the methodology associated with the reference test eg, skill of the pathologist.^41,46^

### Limitations

A comprehensive literature search was conducted and a meticulous screening process was performed. In order to reduce the risk of publication bias, we did not implement any language or publication status restriction. However, it was not possible to search gray literature. The possibility of unpublished studies always exists. The potential for publication bias was not assessed due to the lack of validated methods for diagnostic test accuracy reviews.

## Conclusions

In conclusion, pre-operative endometrial sampling, particularly using Pipelle or conventional curettage, is accurate in diagnosing endometrial cancer. There is insufficient high quality evidence regarding the diagnostic accuracy of other endometrial sampling tests.

## Data Availability

All data produced in the present work are contained in the manuscript

https://osf.io/fp3st/

## Acknowledgments

We acknowledge the effort and the spirit of goodwill of Farida ElShafeey, Menna Kamel, and Aya Ramadan for their contribution to title and abstract screening.

## Author contributions

AN conceived the idea for this review and designed the review methods.

NS, ME, SL collaborated in searching, screening, selecting studies, data extraction and synthesis.

AN wrote the first draft of the manuscript and NS, ME, AZ, MS, AN reviewed the manuscript.

All authors read and approved the final version of the manuscript.

## Availability of data and materials

All data generated or analyzed during this study are included in the published scoping review article and are available in the project public repository.

## Ethics approval and consent to participate

Not applicable

## Consent for publication

Not applicable

